# Beyond the limits of circadian entrainment: Non-24-hour sleep-wake disorder, shift work, and social jet lag

**DOI:** 10.1101/2021.10.06.21264588

**Authors:** Casey O. Diekman, Amitabha Bose

## Abstract

While the vast majority of humans are able to entrain their circadian rhythm to the 24-hour light-dark cycle, there are numerous individuals who are not able to do so due to disease or societal reasons. We use computational and mathematical methods to analyze a well-established model of human circadian rhythms to address cases where individuals do not entrain to the 24-hour light-dark cycle, leading to misalignment of their circadian phase. For each case, we provide a mathematically justified strategy for how to minimize circadian misalignment. In the case of non-24-hour sleep-wake disorder, we show why appropriately timed bright light therapy induces entrainment. With regard to shift work, we explain why reentrainment times following transitions between day and night shifts are asymmetric, and how higher light intensity enables unusually rapid reentrainment after certain transitions. Finally, with regard to teenagers who engage in compensatory catch-up sleep on weekends, we propose a rule of thumb for sleep and wake onset times that minimizes circadian misalignment due to this type of social jet lag. In all cases, the primary mathematical approach involves understanding the dynamics of entrainment maps that measure the phase of the entrained rhythm with respect to the daily onset of lights.

## 1 Introduction

Circadian rhythms are class of endogenous oscillations that occur with a period of roughly 24 hours in a variety of animal and plant species. In humans, circadian oscillations are manifested in a number of ways that range from mRNA and protein levels within cells up to whole body changes in physiology and behavior over the 24-hour cycle [1]. A critical feature of a circadian oscillator is its ability to become phase-locked to periodic environmental signals, such as light and temperature, in a process called entrainment [2, 3]. In mammals, neurons within the suprachiasmatic nucleus (SCN) form the central circadian clock, receiving light-dark input via the retinohypothalamic tract and producing rhythmic output that regulates circadian rhythms throughout the brain and body. A well-studied marker of the human circadian clock is core body temperature (CBT) which oscillates over the course of a full day. The local maximum of CBT usually occurs in mid-afternoon, while the local minimum, CBT_min_, typically occurs in the early morning hours near the end of sleep.

Typically, a person’s circadian clock is entrained to a 24-hour periodic light-dark (LD) cycle. In this paper, we are interested in two distinct situations that deviate from this norm. First, we consider the case of non-24-hour sleep-wake disorder, which afflicts 55-70% of blind people and also can occur in sighted individuals with reduced light sensitivity [4–6]. Individuals with this disorder are unable to entrain to the normal 24-hour periodic LD signal. Instead, they experience a free-running sleep-wake rhythm such that the timing of their sleep progressively shifts each day according to the period of their intrinsic circadian clock (*τ*). Specifically, for *τ* > 24 h, sleep times slowly drift later and later each day, whereas for *τ* < 24 h sleep drifts earlier and earlier. This results in a cyclic sleep disorder with waves of nighttime insomnia and daytime sleepiness relapsing and remitting over weeks and months [7, 8].

We then consider cases in which the LD input either changes abruptly due to night shift work or is not the same over each day of the week due to changes in sleep timings on the weekends. The circadian misalignment that results from transitioning from day shift to night shift leads to decreased alertness and performance levels compared to entrained night shift workers [9]. Furthermore, rotating or permanent night shift workers exhibit increased incidence of cardiovascular disease and cancer relative to permanent day shift workers [10]. The circadian misalignment caused by a change in sleep timings on the weekends, referred to as social jet lag, is also associated with negative behavioral and health outcomes, such as worse academic performance, higher levels of aggression, risk for metabolic disorders and obesity, and depressive symptoms [11].

To study cases that are beyond the normal limits of circadian entrainment, we utilize a limit-cycle model of the human circadian pacemaker due to Forger, Jewett and Kronauer (FJK) [12]. The FJK model is based on experiments measuring the response of the human circadian clock to changes in light exposure and has become a widely-used tool for predicting circadian phase. The model has been extensively validated over a range of laboratory and field conditions [13–16]. To assess entrainment and the properties that lead to it, we will extend and generalize our earlier work on entrainment maps [17, 18]. The entrainment map tracks the position of the entraining oscillator relative to the phase of light onset on a cycle-by-cycle basis as a function of intrinsic period (normal considered to be 24.2 hours) and photoperiod. We have previously used the map with a single light intensity to study reentrainment and jet lag due to rapid switches of time zones [18]. Here we extend the map to consider more realistic multi-lux scenarios in which the intensity of light varies throughout the day. Using a combination of direct simulations of the FJK model and entrainment map analysis, we seek to gain better insight into non-entrained behavior. Our goal is to highlight previously unforeseen effects as well as to suggest interventions that may minimize circadian misalignment.

Regarding individuals with non-24-hour sleep-wake disorder, we will show that they can be made to entrain to a 24-hour LD cycle through the use of one hour of appropriately timed bright light therapy. A practical insight from our results is that the time of day at which the bright light therapy is administered is critical to its success. Non-24 patients are often administered light therapy early in the day upon waking [4, 8, 19]. Our results confirm that such timing can induce entrainment, but only for individuals who have a slower than normal intrinsic body clock. For those with a faster than normal clock, light therapy early in the day fails. Instead, our results show that the therapy will be successful if administered in the evening.

Concerning shift work, when workers change from a normal day time shift to a night time one (or vice versa), they must reentrain to the light-dark protocol of the new shift. This entrainment process can take several days during which the danger of excessive sleepiness during work or driving hours exists. Assuming that there are three possible work shifts (normal day time, late afternoon/evening, and overnight), we use the entrainment map to calculate the reentrainment time as workers change between these three different shifts. We show two key results. First, the reentrainment times due to a transition between any two shifts are asymmetric, similar to the east-west asymmetry in jet lag. For example, the reentrainment time for a switch from the day time to overnight shift is different than the switch from the overnight to daytime shift. At low light intensity, these times can be quite long. Second, with high light intensity, the reentrainment time after certain switches between shifts can be dramatically reduced. This reduction in time is predicted by the entrainment map and is related to a mathematical object called the “phaseless set” [20]. One goal of the analysis is to raise awareness of situations in which the reentrainment time can be reduced, thereby leading to strategies to enhance this possibility.

Social jet lag can occur when an individual changes their sleep timing for a few days per week, typically over the weekend, thereby changing their light exposure on those days. In this case, since the LD input is not 24-hour periodic, the circadian oscillator does not entrain to a 24-hour cycle. Instead, we show that the oscillator entrains to a complicated seven-day pattern in which the circadian oscillator is misaligned for part of the week. We illustrate this using the example of teenagers who sleep too little during the school week and try to compensate by sleeping more over the weekend. We show that the timing (sleep and wake time) of this compensatory sleep is critical to reducing misalignment and that simply sleeping more independent of the timing is not an optimal strategy to combat misalignment.

## 2 Methods

The FJK model [12] describes the circadian variation of core body temperature and is based on prior models of Kronauer and collaborators [21–23]:

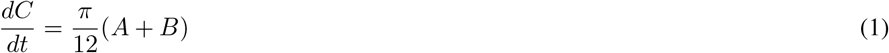

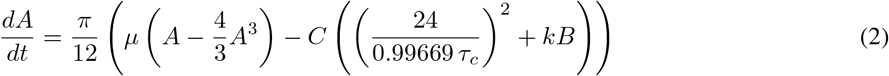

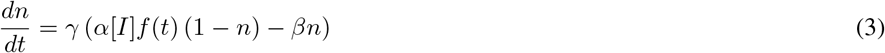

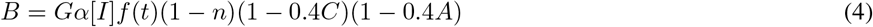

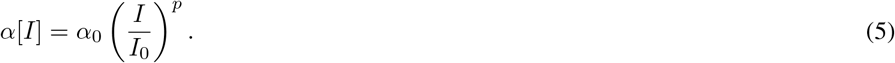

The variable *C* represents core body temperature, *A* is a phenomenological auxiliary variable, and *n* models the phototransduction pathway through which light drives the circadian system. The variable *B* models circadian modulation of the oscillator’s sensitivity to light. All parameter values are positive. The parameter *τ*_*c*_ determines the period of the oscillator in constant darkness, *I* codes for the intensity of light, *G* is a gain factor that determines the extent of light that actually reaches the SCN, and *p* controls the steepness of the dose-response curve to light. The parameters *μ* = 0.23, *k* = 0.55, *γ* = 60, *β* = 0.0075, *α*_0_ = 0.05, *I*_0_ = 9500 and *G* = 33.75 are kept fixed throughout this study. In addition, we choose *τ*_*c*_ = 24.2 and *p* = 0.5 to represent an individual with a normal clock and dose-response factor. To model constant light, we take *f* (*t*) ≡ 1, whereas constant darkness occurs with *f* (*t*) ≡ 0. In the FJK model, both situations lead to periodic solutions referred to as the LL and DD limit cycles, respectively.

We shall consider a variety of light/dark (LD) protocols by taking *f* (*t*) to be a square wave with a period of 24 hours and a duty cycle of *N/*24 where *N* is the photoperiod, i.e. *f* (*t*) ≡ 1 for *N* hours of light and *f* (*t*) ≡ 0 for − 24 *N* hours of darkness. As a base case, we shall consider days with *N* = 17 hours of light exposure in which the light intensity varies depending on the time of the day as well as whether the individual is assumed to be indoors or outdoors. As a canonical case, we consider sunrise at 7AM with sunset at 7PM meant to mimic the spring or fall equinox. Individuals wake up at 6AM and experience one hour of indoor light at *I* = 100 lux. This is followed by 12 hours of natural sunlight with *I* = 1000 lux and 4 hours of indoor light at *I* = 100. Sleep occurs from 11PM until 6AM in complete darkness. We use the notation *N* = 17 : 1(12)4, *I* = 100(1000)100 to denote this light pattern and will suitably adapt it for other scenarios.

In [17], we developed an entrainment map to assess whether and at what phase a circadian oscillator entrains to a periodic LD forcing with a single light intensity. The entrainment map records the number of hours that have passed since the lights last turned on each time the trajectory returns to a pre-chosen Poincaré section of the phase space. For example, choose the Poincaré section 𝒫, at *A* = 0, with *A*′ > 0, which yields a rectangle in the *C* and *n* space. Assume that an oscillator has an initial condition that lies on 𝒫 with *n* = 0 and the *C* value chosen as the value at the intersection of 𝒫 with the DD limit cycle. Let *x* denote the number of hours since the lights last turned on. Evolve the trajectory under the flow until it again returns to 𝒫. Call this time *ρ*(*x*). The entrainment map Π(*x*) is defined as the amount of time that has passed since the most recent onset of the lights. In [17], we showed that Π(*x*) = (*x* + *ρ*(*x*)) mod 24, which yields a one-dimensional map. The entrainment map is periodic in that Π(0^+^) = Π(24^−^) and maps the interval [0,24] onto itself. Typically, it has at most one point of discontinuity and is increasing otherwise. It depends continuously on parameters of interest such as *τ*_*c*_, *p*, and *I*. In many cases, including those considered in this work, a stable fixed point of the entrainment map can be related to the existence of a stable entrained circadian rhythm of exactly 24 hours.

The one-dimensional entrainment map Π can have two, one, or zero fixed points. When it has two fixed points, one of them is stable and denoted *x*_*s*_, while the other is unstable and denoted *x*_*u*_. The situation that corresponds to the existence of one fixed point occurs when varying a parameter causes *x*_*s*_ and *x*_*u*_ to merge at a saddle-node bifurcation. A further change of this parameter causes there to be no fixed points. When *x*_*s*_ exists, we say that the circadian oscillator is entrained to the 24-hour LD forcing and call it an LD-entrained solution.

In the first two parts of this study, we shall be interested in cases where we can utilize a single multi-lux entrainment map to study properties of entrainment. These cases arise when an individual follows the same light-dark schedule every day over several weeks. For the case of non-24-hour sleep-wake disorder, we will show how to induce a stable fixed point of the map, and thus entrainment, via bright light therapy. For the case of shift workers, we will investigate the transient dynamics determining how long it takes a shift worker to reentrain after a switch from one shift to another (e.g. normal day time shift to a night shift).

In the third part of our study, we consider the case of social jet lag where individuals are exposed to different light schedules on different days of the week, although we shall assume that the light schedules follow a repeating seven-day pattern. In these cases, there are multiple multi-lux entrainments maps that need to be utilized over the course of one week. While each of these maps has a stable fixed point, the trajectory never converges to either fixed point because of the differing amounts of light exposure on different days. Instead, we will show that the trajectory converges to a periodic orbit which can be found through a generalized cobweb procedure. In these cases, we say that the individual is entrained to a 7-day cycle. Here we will be concerned about the extent of misalignment of the circadian oscillator from the stable 24-hour LD-entrained solution that would occur in the absence of social jet lag.

Entrainment maps were computed numerically through simulations of the FJK model using the *ode15s* solver in MATLAB. When cobwebbing the map to model the reentrainment process, an orbit is considered entrained when the iterates converge to within 0.5 hours of the stable fixed point of the map. For direct simulations of the FJK model, we define a trajectory to be entrained when it crosses a 1D section of the phase space (e.g., *A* = 0 with *A*′ > 0 as in Fig. 1) within 0.5 hours of when a reference trajectory that has already converged to the stable limit cycle for the current LD protocol (i.e. an LD-entrained periodic solution) crosses the section.

**Figure 1:**
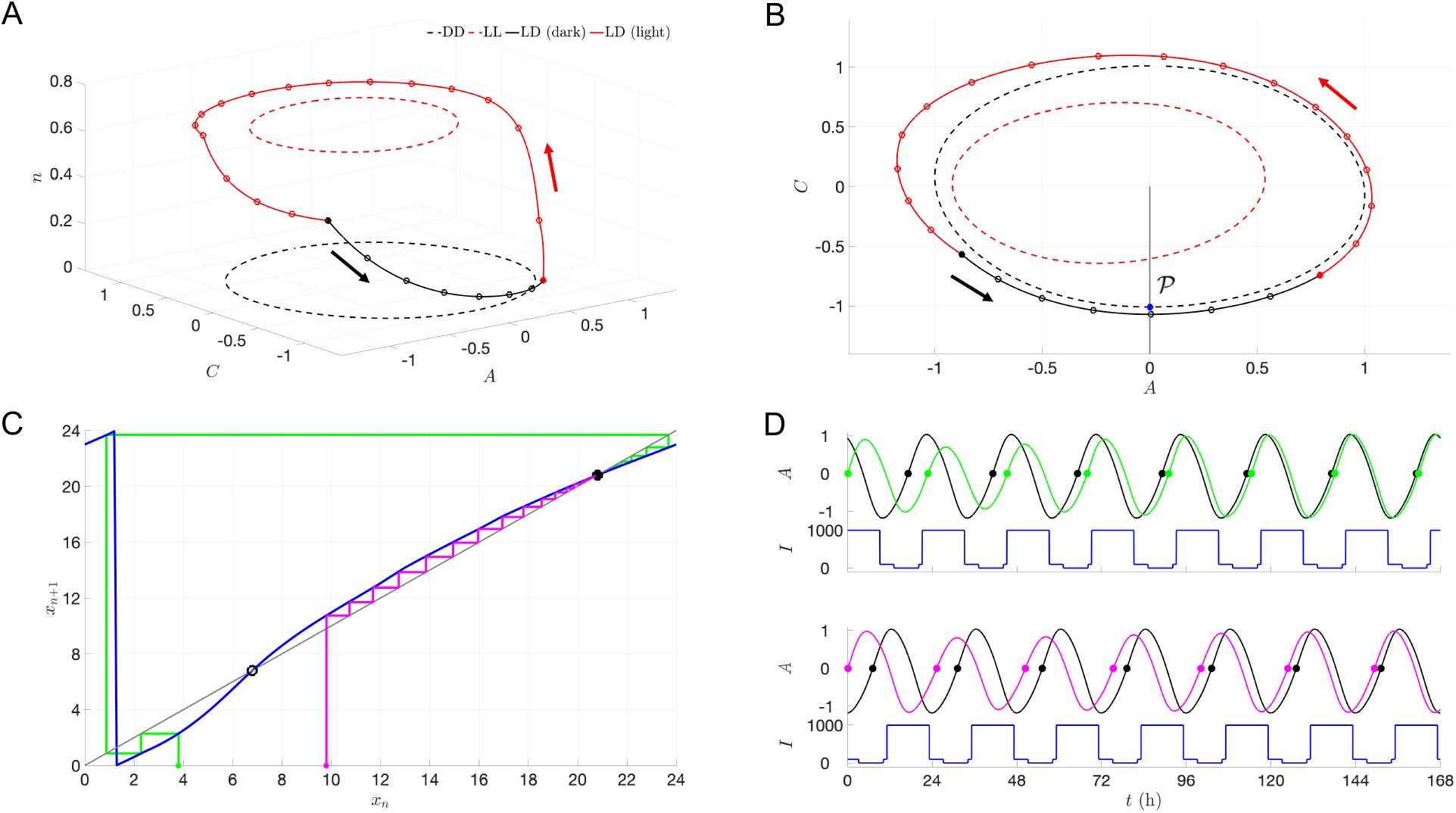
Limit cycles, entrainment map, and reentrainment simulations for the FJK model. Parameter values: *τ*_*c*_ = 24.2, *p* = 0.5, *N* = 1(12)4, *I* = 100(1000)100. *A:* DD (*I* = 0, dashed black), LL (*I* = 1000, dashed red), and LD-entrained (solid black and red) limit cycles in the *A* − *C* − *n* phase space. Hourly markers with lights on (starting at solid red dot) and lights off (starting at solid black dot). Red and black arrows depict the counterclockwise direction of the flow. *B:* Projection of limit cycles in (A) onto *A* − *C* phase plane. Poincaré section 𝒫 (gray line) at *A* = 0 with *A*′ *>* 0 and its intersection with the DD limit cycle (blue dot). *C:* Entrainment map computed with section 𝒫 and initial conditions (blue dot) shown in (B). Stable fixed point *x*_*s*_ (solid black dot), unstable fixed point *x*_*u*_ (open black dot), and map iterates converging to *x*_*s*_ through phase advances (green) and phase delays (magenta). *D:* Simulations of FJK model with trajectories converging to an entrained reference trajectory (black curves) through phase advances (top panel, green curve) and phase delays (bottom panel, magenta curve) following a shift of the LD cycle (blue curves). The solid dots along each time course provide references corresponding to when the relevant trajectory crosses the Poincaré section, allowing for easy comparisons of phase.

## 3 Results

This section contains three specific studies. The first study is on non-24-hour sleep-wake disorder and explores how the existence of an entrained solution depends on various parameters by showing circumstances under which the multi-lux entrainment map gains or loses a fixed point. The second study is on shift workers and examines the reentrainment time when a worker switches between different work shifts. We initially consider scenarios with single-lux maps of different light intensities and then move on to scenarios involving pairs of multi-lux maps. The third study is on social jet lag and addresses the degree of misalignment when individuals follow a periodic seven day light-dark schedule that varies from day-to-day. For this application, we utilize several entrainment maps and a generalized cobweb procedure. In all applications, we use entrainment maps in conjunction with direct simulations to explain the underlying dynamics.

In Fig. 1, we illustrate how the entrainment map works for a multi-lux case, *N* = 1(12)4, *I* = 100(1000)100. Figure 1A shows the LD-entrained limit cycle (solid red and black curve) in the *A* − *C* − *n* phase space. Hourly marking are demarcated, where the red portion of the curve signifies where lights are on and the black part where lights are off. Note the dip in the trajectory to an intermediate *n* value for the hours corresponding to 7PM-11PM when the light intensity changes from 1000 to 100 lux. The dashed red and black limit cycles lie in constant *n* planes and signify the LL and DD solutions, respectively. Figure 1B shows the projection of those limit cycle solutions on a common *A* − *C* phase plane, together with the Poincaré section at *A* = 0, *A*′ > 0. Figure 1C shows the 1-dim entrainment map (blue curve). The map is 24-hour periodic and exhibits a discontinuity at *x* = 1.25 due to the mod 24 operation. On intervals where it is continuous, the map is increasing. Note that the map intersects the diagonal (gray) line at two points representing a stable (solid black dot) and an unstable (open black dot) fixed point. Additionally, we show cobwebs from two distinct initial conditions leading to solutions that converge to the stable fixed point by either phase advance (green) or delay (magenta). A key point is that the unstable fixed point splits the *x*-axis into regions from where solutions converge via either advance or delay. Figure 1D shows the direct simulation of these phase advancing and delaying solutions on their path to convergence to the LD-entrained solution (black curve).

### 3.1 Non-24-Hour Sleep-Wake Disorder

To develop a basis for therapeutic light therapy to counteract non-24-hour sleep-wake disorder, we shall make two assumptions. First, we assume that the individual has an intrinsic period that differs from the norm of 24.2 hours. The intrinsic period can either be long (24.7 hours) or short (23.5 hours). Second, we shall assume that such individuals have a reduced sensitivity to light, which we model by increasing the value of the dose-response exponent *p* from 0.5 to 1.5. These assumptions are consistent with a recent study on interindividual variability in circadian timing [24]. The goal of our analysis will be to determine whether one hour of appropriately timed bright light therapy can induce a stable fixed point of the map that corresponds to CBT_min_ occurring during sleep.

In Fig. 2, we illustrate how the multi-lux entrainment map depends on the parameters *τ*_*c*_ and *p*. In Fig. 2A, we keep the dose-response exponent *p* = 0.5, while varying *τ*_*c*_. As *τ*_*c*_ increases, the intrinsic rate of evolution of the oscillator decreases, so it takes longer for a trajectory to return to the Poincaré section. Thus the entrainment map shifts up. Alternatively, decreasing *τ*_*c*_ shifts the map down. In Fig. 2B, the intrinsic rate *τ*_*c*_ = 24.2 is constant, but the dose response exponent is changed. As *p* increases, the *α*[*I*] term in equation (5) decreases (since *I* < *I*_0_). This has the effect of dampening the effect of light, and the entrainment map flattens out. In fact, as *p → ∞*, the effect of the LD forcing would disappear and the entrainment map would become a line with constant slope, lying above (below) the diagonal when *τ*_*c*_ > 24 (*τ*_*c*_ < 24). In Fig. 2C, we show the effect of varying *τ*_*c*_ and *p* simultaneously. With *p* = 1.5, the maps no longer have fixed points for either *τ*_*c*_ = 23.5 or 24.7. Instead, the maps lie below or above the diagonal. These cases corresponds to non-24-hour sleep-wake disorder. Finally, in Fig. 2D, we show how the non-entrainment manifests itself for *p* = 1.5 by plotting the time course of *C* (which corresponds to the CBT) in direct simulations. The oscillation in the envelope of the time course indicates non-entrainment to the LD-cycle and instead convergence to a longer-period cycle that stretches over tens of days.

**Figure 2:**
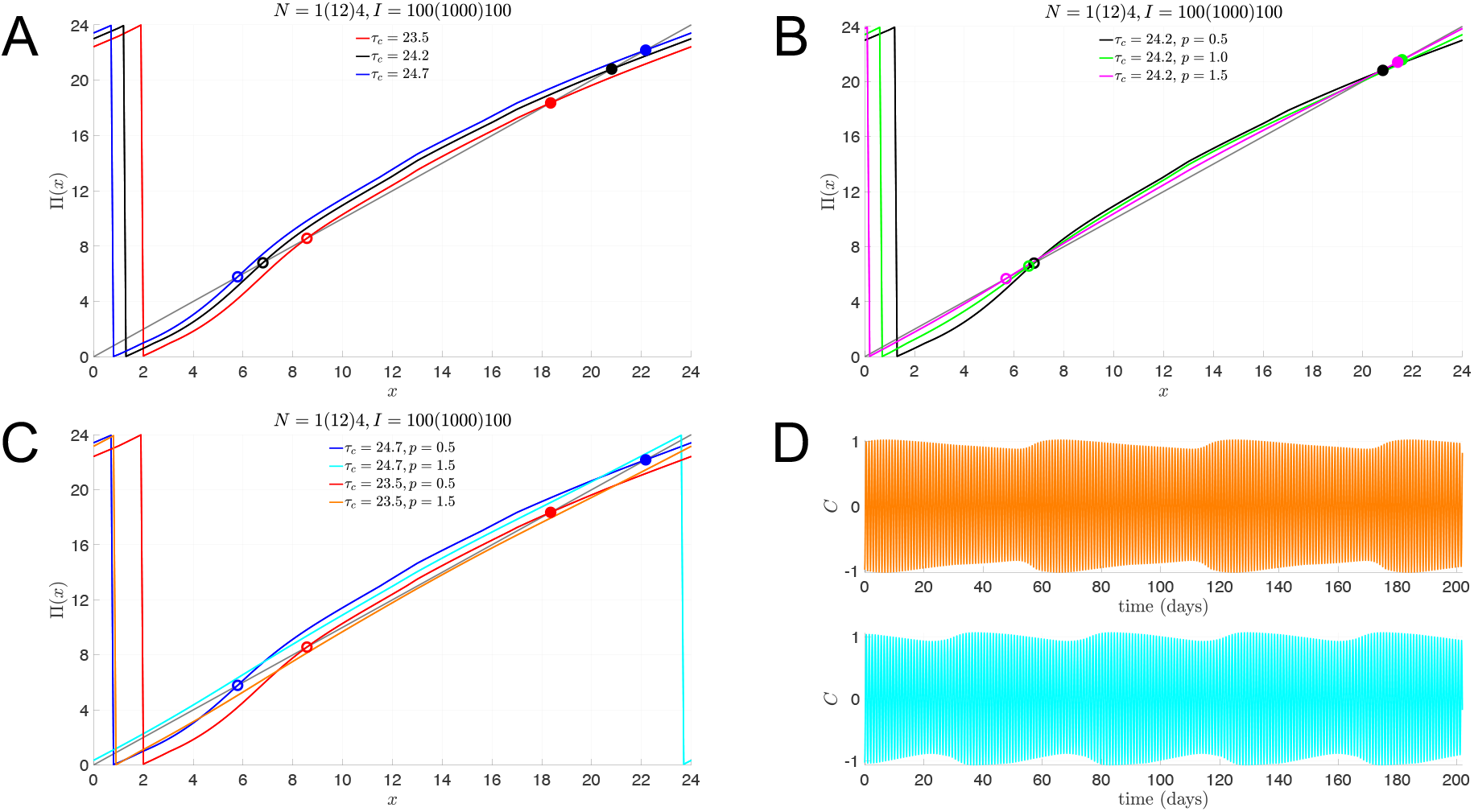
Loss of entrainment with slow or fast intrinsic clocks and reduced light sensitivity. Total light duration of *N* = 17 hours, composed of 1 hour with *I* = 100, 12 hours with *I* = 1000, and 4 hours with *I* = 100. *A:* Entrainment maps for slow (*τ*_*c*_ = 24.7, blue), normal (*τ*_*c*_ = 24.2, black), and fast (*τ*_*c*_ = 23.5, red) intrinsic clocks with normal light sensitivity (*p* = 0.5) have stable fixed points (solid dots) representing the stable phase of entrainment of CBT_*min*_. *B:* Entrainment maps for normal (*p* = 0.5, black) and reduced (*p* = 1.0, green; *p* = 1.5, magenta) light sensitivity with normal intrinsic clocks (*τ*_*c*_ = 24.2, black) also have stable fixed points (solid dots) representing the stable phase of entrainment. *C:* Entrainment maps with reduced light sensitivity (*p* = 1.5) and slow (*τ*_*c*_ = 24.7, cyan) or fast (*τ*_*c*_ = 23.5, orange) intrinsic clocks do not have stable fixed points. *D:* Time course of *C* variable from simulations of the full model showing the lack of 1:1 entrained solutions for parameter values corresponding to the maps without stable fixed points (orange: *τ*_*c*_ = 23.5, *p* = 1.5; cyan: *τ*_*c*_ = 24.7, *p* = 1.5).

That the *p* = 1.5 entrainment maps for *τ*_*c*_ = 23.5 and 24.7 no longer have fixed points is illustrated by computing a cobweb diagram for these cases (Fig. 3A, B). The cobweb diagrams reveal that CBT_min_ systematically phase advances (delays) over weeks for each of these cases. The period of these orbits is 61 (44) days for *τ*_*c*_ = 23.5 (24.7), the length of which is directly related to how close the entrainment map lies to the diagonal. To explore whether the introduction of bright-light therapy can induce LD entrainment, we introduce one hour of 10, 000 lux light at various points of the day and recompute the entrainment maps. The idea that high intensity light may aid in the entrainment process is supported by two intertwined mathematical reasons. First, when *I* > *I*_0_ = 9, 500, then *α*[*I*] > 1, which would tend to increase the effect of light. Second, from our earlier studies [17, 18], increased light intensity increases the concavity of the entrainment map. In combination, these two effects may be enough to induce a fixed point in the entrainment map, and thus LD-entrainment. In Panels C and D, we show the effect of adding one hour of bright light immediately upon awaking, from 6AM to 7AM. The corresponding maps, *N* = 1(12)4, *I* = 10, 000(1000)100, do exhibit greater concavity, most pronouncedly for small values of *x*. The effect is to create a pair of fixed points. Importantly, the stable fixed point associated with *τ*_*c*_ = 24.7 occurs at *x* = 21.4, which corresponds to roughly 3:30AM. As noted earlier, this stable fixed point corresponds to the phase (and time) at which the CBT_min_ occurs. Thus, early morning bright light therapy not only induces a stable LD-entrained solution for those with a slow body clock, but also does so with a phase that is well aligned to the norm. However, when an individual’s body clock is too fast, *τ*_*c*_ = 23.5, the LD-entrainment that does occur (Panel D) yields a stable phase that falls in the middle of the day at *x* = 8.5. Thus, the CBT_min_ value does not occur during the night as would be desired. To attempt to correct for this misalignment, we checked the effect of one hour of bright light therapy in the early evening, from 7PM-8PM (Panels E and F). The single hour of high intensity light in the evening has the therapeutic effect of inducing a stable fixed point at *x* = 20.5, which falls squarely within the night-time sleep period. For the case of *τ*_*c*_ = 24.7, however, the bright light therapy in the evening does not create a fixed point. In fact, the cycle period increases to over 100 days, and most days are spent with a CBT_min_ value falling during waking hours. Thus, bright light therapy for those with a faster than normal body clock should be given in the evening, not the morning; for those with longer than normal body clocks, bright light therapy in the morning is effective, but is detrimental if given in the evening.

**Figure 3:**
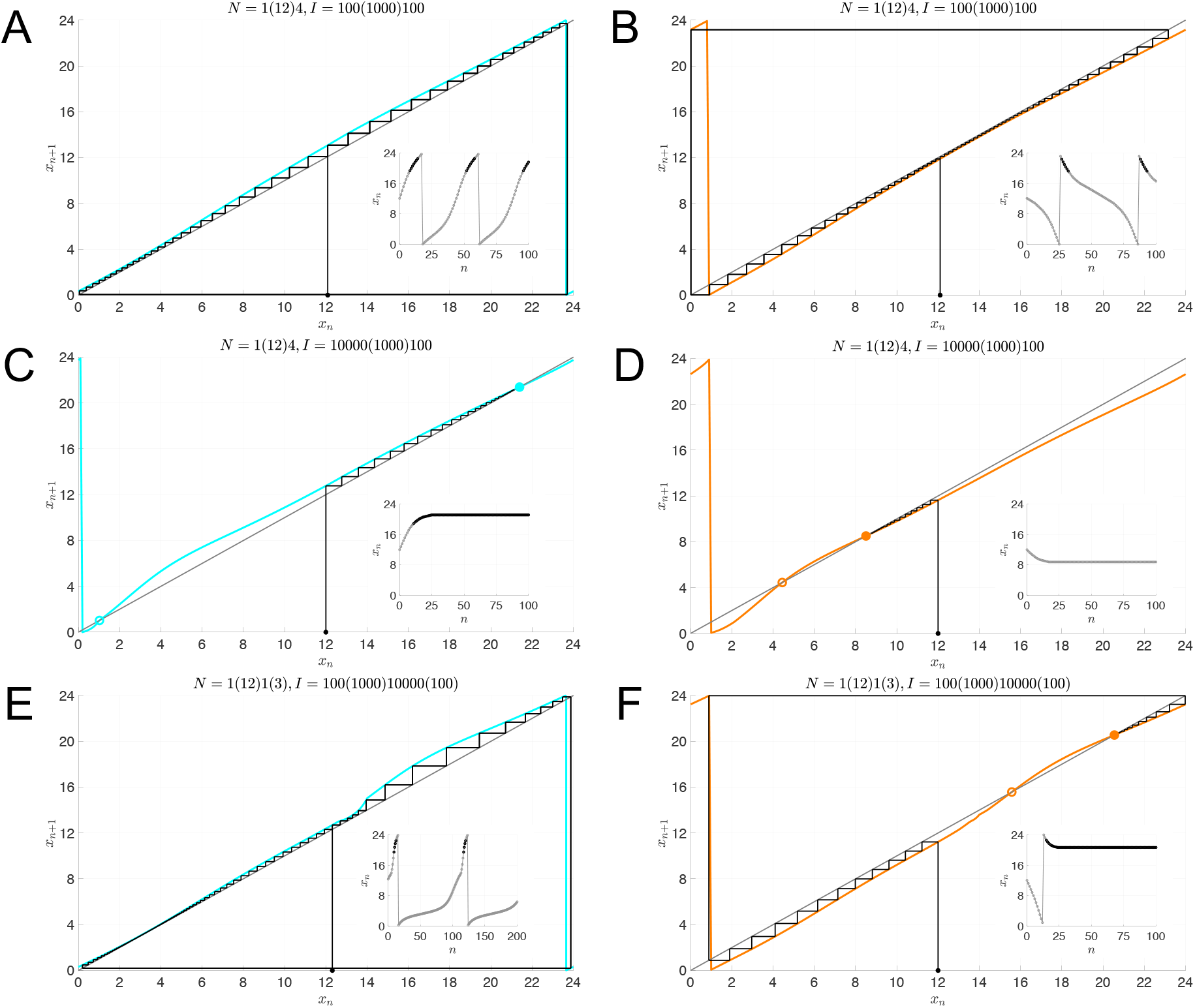
Entrainment map analysis of non-24-hour sleep-wake disorder and bright light therapy. *A-F:* Cobweb diagrams for entrainment maps with *p* = 1.5 and *τ*_*c*_ = 24.7 (cyan) or *τ*_*c*_ = 23.5 (orange) starting with an initial condition of *x*_0_ = 12 (solid black dot). Insets show the iterates *x*_*n*_ as a function of *n*; black iterates are those within 2 hours of *x* = 20.8, which is the stable phase of entrainment for the normal situation of *τ*_*c*_ = 24.2 with *N* = 1(12)4 and *I* = 100(1000)100 as shown in Fig. 1C. *A-B:* Maps for a total light duration of *N* = 17 hours, composed of 1 hour with *I* = 100, 12 hours with *I* = 1000, and 4 hours with *I* = 100. There is no stable fixed point and the iterates converge to periodic orbits. *A:* Orbit for *τ*_*c*_ = 24.7 with continual phase delays and a period of 44 days. *B:* Orbit for *τ*_*c*_ = 23.5 with continual phase advances and a period of 61 days. *C-D:* One hour of bright light (10, 000 lux) administered first thing in the morning induces stable fixed points in both maps. *C:* Phase of entrainment for CBT_min_ is in the evening (*x*_*s*_ = 21.4) for *τ*_*c*_ = 24.7. *D:* Phase of entrainment for CBT_min_ is in the afternoon (*x*_*s*_ = 8.5) for *τ*_*c*_ = 23.5. *E:* One hour of bright light (10, 000 lux) administered in the evening does not induce a stable fixed point for *τ*_*c*_ = 24.7. The orbit exhibits continual phase delays with a period of 108 days. *F:* One hour of bright light (10, 000 lux) administered in the evening induces a stable fixed point for *τ*_*c*_ = 24.7 with a phase of entrainment for CBT_min_ in the evening (*x*_*s*_ = 20.5).

### 3.2 Shift work

We now turn to reentrainment of shift workers when they change their work schedule from one shift to another. Consider the following three-shift scenario: Shift 1 works from 7AM-3PM and sleeps 11PM-6AM; Shift 2 works from 3PM-11PM and sleeps from 12AM-7AM; and Shift 3 works from 11PM-7AM and sleeps 8AM-3PM. We consider a worker on a 7-day work schedule who abruptly transitions from one shift to another. Transitioning from 1st to 2nd, 1st to 3rd, and 2nd to 3rd (or vice versa) are 1-hour, 9-hour, and 8-hour shifts of the sleep-wake schedule. We initially consider two separate *N* = 17, single-lux cases of *I* = 100 or 1000 and study the reentrainment time when a worker changes from working on one of these shifts to any other. Importantly, in these single-lux cases, the limit-cycle solutions for each shift are identical. The only difference is that the clock time associated with points on the limit cycle differ depending on the shift. Thus, we can consider a single entrainment map and assess reentrainment times by varying the initial condition. We then study the more realistic scenario of a multi-lux light protocol. In particular, we assume that normal indoor light is 100 lux, outdoor light is 1000 lux, and light intensity at work is 500 lux. We continue to use *N* = 17 maps but now individuals living on different shift work schedules will be exposed to different light schedules. In turn, this will cause there to be distinct limit cycles for each shift. Thus while considering transitions, we not only need to vary initial conditions, but also the map which is used to obtain the cobweb. In both single and multi-lux cases, we assume the sun rises at 7AM and sets at 7PM.

For the single-lux maps, when calculating reentrainment times for these transitions we take the Poincaré section at the start of sleep for the shift they were previously on (corresponding to 11PM, 12AM, and 8AM for 1st, 2nd, and 3rd shift, respectively); see Fig. 4. Thus the stable fixed point is always at *X* = 17. For the multi-lux maps, the location of the Poincaré section is still the start of sleep at *X* = 17 on the previous shift. However, this section intersects the limit cycle of the new shift at an hourly location that is different than *X* = 17. For example, for transitions between 1st and 3rd shifts, we build the entrainment map based on the hourly marking of the new shift (see Fig. 4C, D). The initial conditions for cobwebbing are based on how many more hours of light the worker will receive on their new shift. For example, when transitioning from 1st to 3rd shift, instead of going to bed at 11PM the worker will receive 9 more hours of light before going to bed at 8AM, so we use *x*_0_ = 8. On the other hand, when transitioning from 3rd to 1st shift, instead of going to bed at 8AM they will receive 15 more hours of light before going to bed at 11PM; thus we use *x*_0_ = 2. Based on similar logic we use *x*_0_ = 16 and *x*_0_ = 18 for 1st to 2nd and 2nd to 1st transitions, and *x*_0_ = 9 and *x*_0_ = 1 for 2nd to 3rd and 3rd to 2nd transitions.

**Figure 4:**
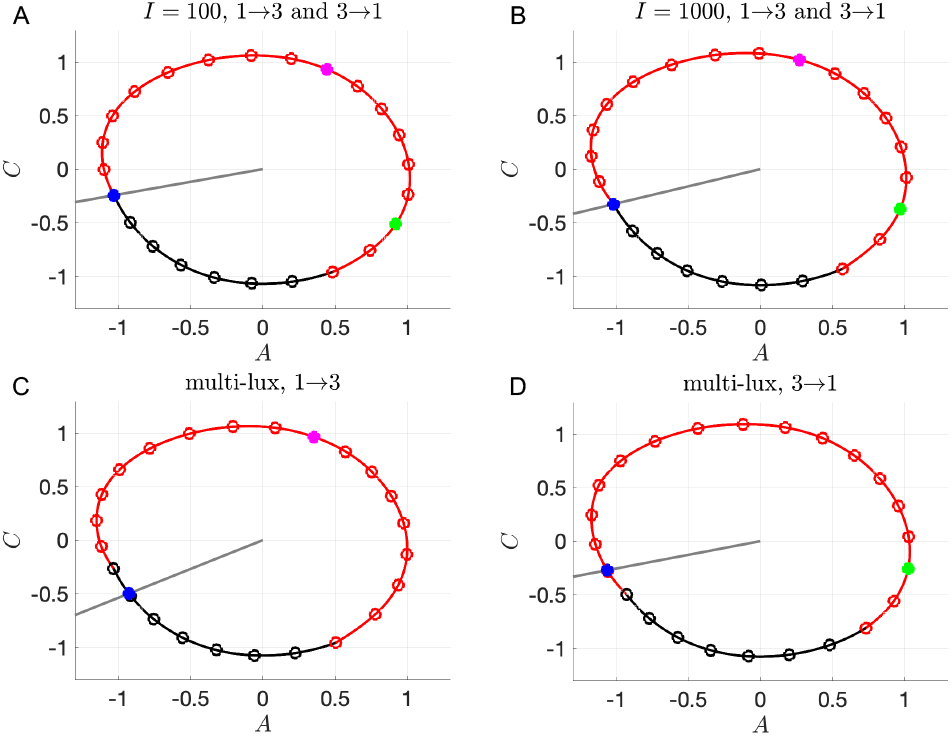
Limit cycles used to compute entrainment maps for transitions between 1st and 3rd shift. Poincaré sections 𝒫 (gray lines), initial conditions for computing the maps (blue dots), locations of entrained 1st shift workers at 8AM (green dots, *X* = 2), and locations of entrained 3rd shift workers at 11PM (magenta dots, *X* = 8). *A-B:* Limit cycle for *N* = 17 single-lux light protocol with *I* = 100 (A) and *I* = 1000 (B). 𝒫 passes through *X* = 17, corresponding to 11PM on 1st shift LC and 8AM on 3rd shift LC. *C-D:* Limit cycles for multi-lux light protocol with *N* = 1(12)4 and *I* = 100(1000)100, with 𝒫 passing through *X* = 17.91 on 3rd shift LC (C) and *X* = 15.96 on 1st shift LC (D).

The bar graph in Fig. 5 shows the reentrainment times for a given pair of transitions (e.g. 1→3 and 3→1) together on the same vertical scale for the different light protocols (single-lux *I* = 100, single-lux *I* = 1000, and multi-lux). As might be expected, the reentrainment for the 1→2 and 2→1 transitions are relatively fast and symmetric. Higher intensity light leads to faster entrainment (compare *I* = 100 to *I* = 1000) due to increased concavity of the single-lux map. Since there is nothing surprising about these transitions we do not show any further results for these cases.

**Figure 5:**
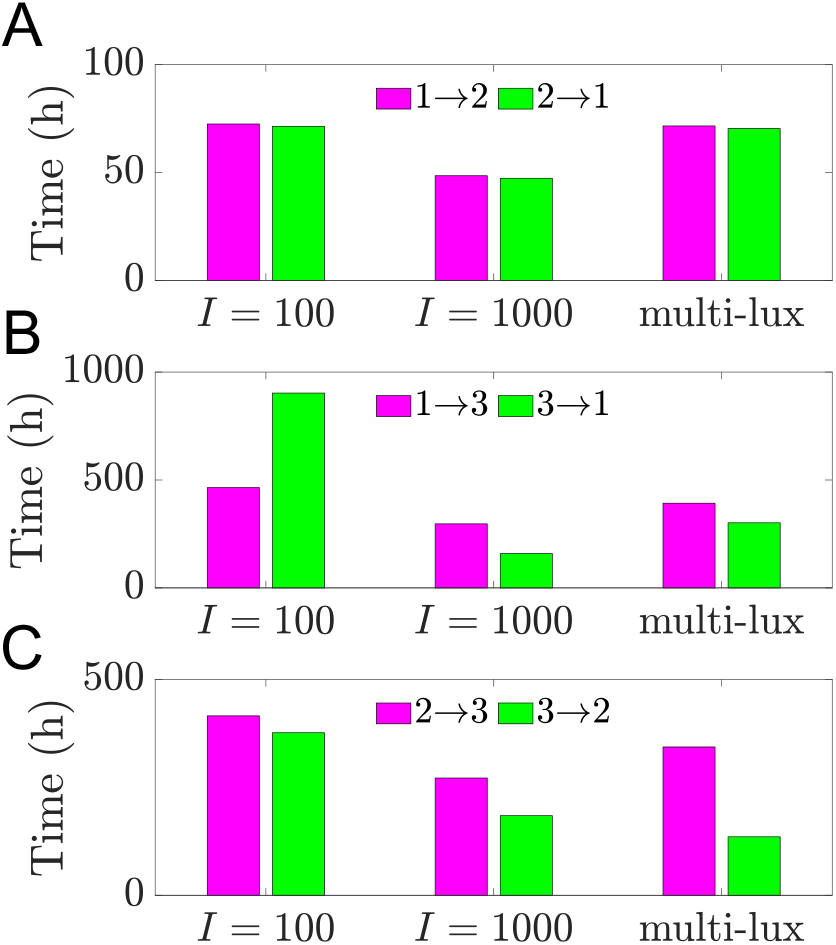
Reentrainment times following transitions between 1st, 2nd, and 3rd shift schedules. Direct simulations of shift work transitions under single-lux (*I* = 100, *I* = 1000) and multi-lux LD protocols. *A:* Transitions from 1st to 2nd shift (magenta) and from 2nd to 1st shift (green) result in similar reentrainment times under all 3 LD protocols. *B:* Transitions from 1st to 3rd shift (magenta) result in shorter reentrainment times for *I* = 100 but longer reentrainment times for *I* = 1000 and multi-lux protocols compared to the transitions from 3rd to 1st shift (green). Entrainment time for 3rd to 1st with *I* = 1000 is shorter than it is with multi-lux. *C:* Transitions from 2nd to 3rd shift (magenta) result in longer reentrainment times than transitions from 3rd to 2nd shift (green) under all 3 LD protocols. Entrainment time for 3rd to 2nd with *I* = 1000 is longer than it is with multi-lux.

The transitions between Shifts 1 and 3 are more interesting as they display not only asymmetries in the reentrainment times, but also considerable differences depending on the light intensity or protocol. The 100 lux entrainment time for the transition from Shift 1 to 3 is considerably shorter than from Shift 3 to 1. This observation is readily explained using the entrainment map shown in Fig. 6A. The transition from Shift 1 to 3 corresponds to cobwebbing with an initial condition *x*_0_ = 8, while the transition from Shift 3 to 1 corresponds to the initial condition *x*_0_ = 2. For this latter case, the initial condition lies very close to the unstable fixed point of the map. Here, a large number of map iterates (each corresponding to roughly 24 hours) is needed for the trajectory to escape a neighborhood of this unstable fixed point. This contributes to the long entrainment time, which occurs via phase advance. In contrast, for the transition from Shift 1 to 3, the initial condition does not lie near the unstable fixed point and the reentrainment is much faster, occurring now via phase delay. Figures 6B and C show time courses for the CBT variable *C* which confirm this map prediction. When the light intensity is increased to *I* = 1000, the reentrainment time for these transitions significantly decreases, and the reentrainment from Shift 3 to 1 becomes dramatically faster than that from Shift 1 to 3 (see Fig. 6E,F and note the different time scales shown in these figures). This reversal in the asymmetry of the reentrainment times upon increasing the light intensity may seem surprising since the entrainment map for *I* = 1000 looks qualitatively similar to the one for *I* = 100; compare Fig. 6A to D. However, in previous work [18], we have found that at higher lux levels, initial conditions that start in a neighborhood of the unstable fixed point can have very *short* entrainment times. This phenomenon is due the existence of a mathematical object known as “the phaseless set” [20], which provides trajectories access to shortcuts in phase space that dramatically reduce entrainment times. A hallmark of this behavior can be seen in Fig. 6F in the form of amplitude suppression of the green trajectory. In the full 3-dimensional phase space, suppression corresponds to the trajectory evolving near the *n*-axis (where *A* and *C* values are small), for which small changes in the location of the trajectory can correspond to large changes in phase. Specifically, note the large phase advance near *t* = 120 h in Fig. 6F, illustrated by the change in the horizontal position of the green dot compared to the reference black dot.

**Figure 6:**
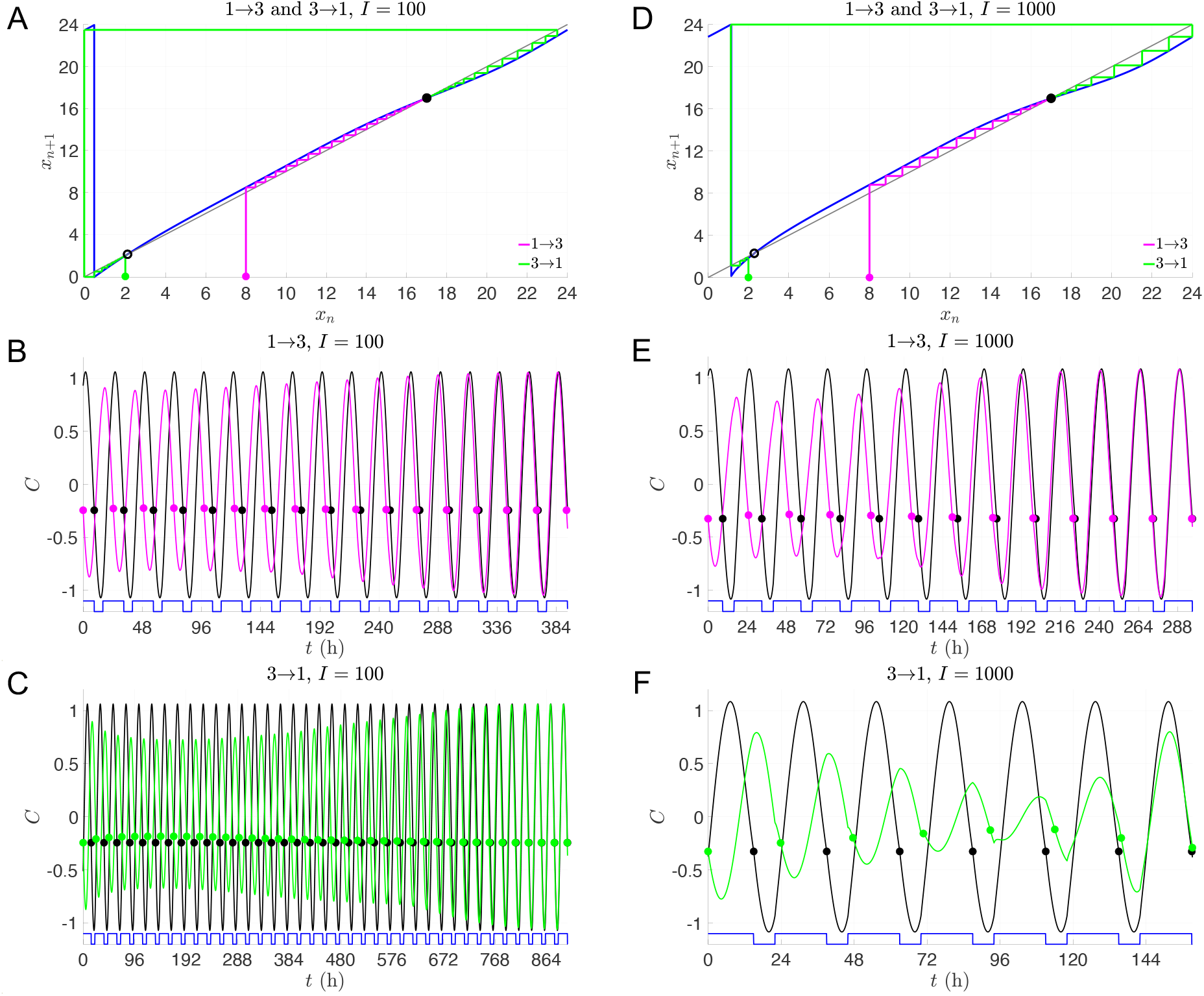
Dynamics of transitions between 1st and 3rd shifts with single-lux LD protocols. *A-C:* Entrainment maps and cobweb diagrams (A) and time courses of direct simulations of 1→3 (B) and 3→1 transitions (C) with *I* = 100. *D-F:* Entrainment maps and cobweb diagrams (D) and time courses of direct simulations of 1→3 (E) and 3→1 transitions (F) with *I* = 1000. Note that a different time scale is shown for each time course.

For the more realistic multi-lux case, the reentrainment times fall in between those of the *I* = 100 and *I* = 1000 lux cases (Fig. 5), though they lie closer to the *I* = 1000 results. The entrainment maps for the multi-lux case, Fig. 7A and B, are qualitatively similar to the single-lux maps. However the Shift 1 to 3 multi-lux map has a flatter region than the *I* = 1000 lux map that lies closer to the diagonal for an interval of *x* values near (4, 8). This flattening causes the entrainment in the multi-lux case to be longer as the initial condition *x*_0_ takes a number of iterates to evolve from there. The transition from Shift 1 to Shift 3 is through phase delay (magenta dots phase recess relative to the black ones for the time course shown in Fig. 7C, consistent with the cobweb diagram in Fig. 7A). The Shift 3 to 1 transition for the multi-lux case displays dramatically shorter entrainment times than the 100 lux case and is closer to the 1000 lux time. This is again due to the phaseless set as the initial value *x*_0_ = 2 lies very close to the unstable fixed point of the map (Fig. 7B). The time course (Fig. 7D) not only displays amplitude suppression, but also another hallmark of the phaseless set in that there is not a consistent direction of entrainment through either phase advance or phase delay. The green dots initially do not change their phase relative to the black. Then suddenly between *t* = 96 and 120 there is a dramatic phase advance, and from *t* = 120 to 144 there is a phase delay, but such that the actual phase is almost completely aligned to the reference phase! Thus the notion of converging to the stable phase of entrainment phase through successive advances or delays is lost.

**Figure 7:**
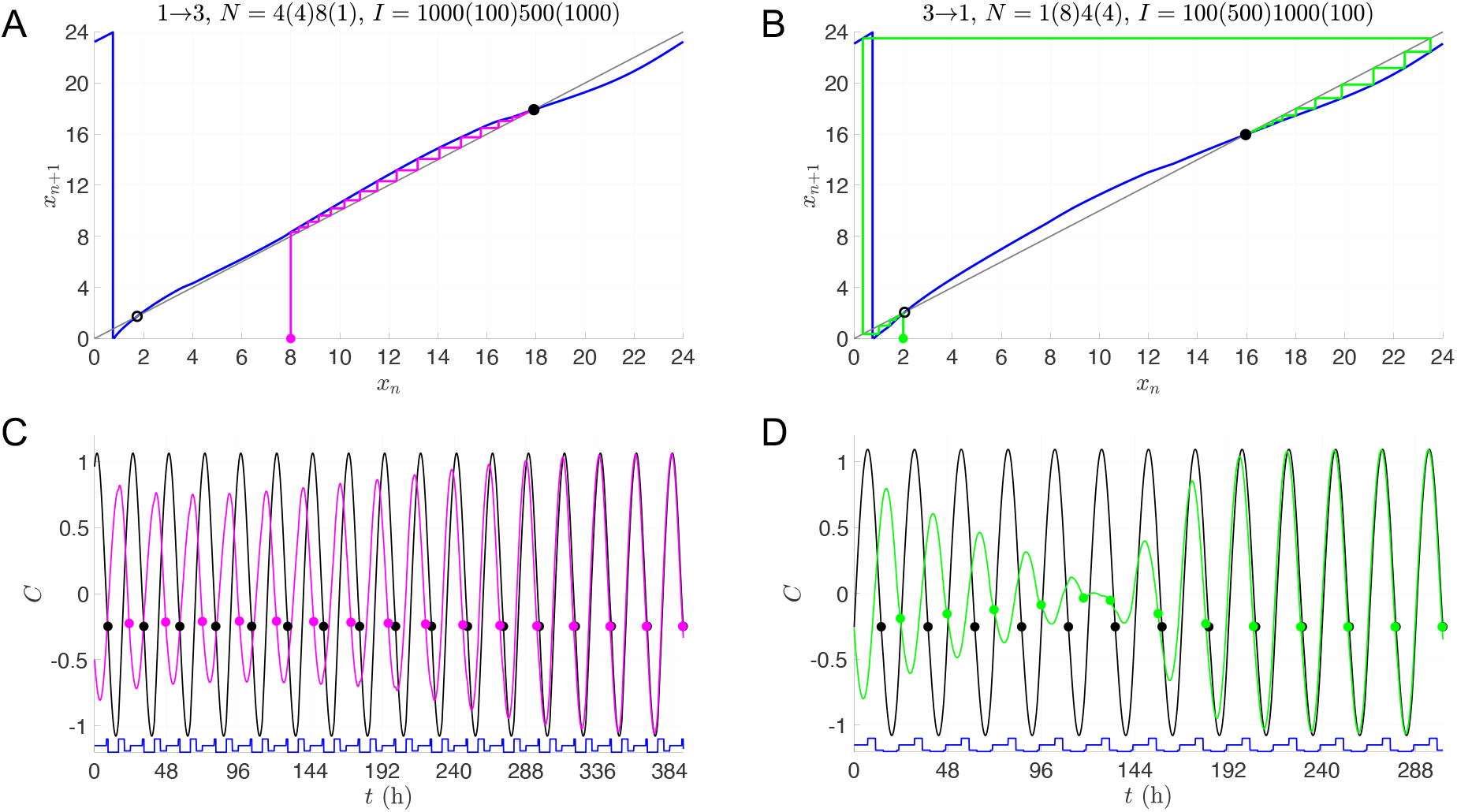
Dynamics of transitions between 1st and 3rd shifts with multi-lux LD protocols. *A,C:* Entrainment maps and cobweb diagrams (A) and time course of direct simulation (C) for 1→3 transition. *B,D:* Entrainment maps and cobweb diagrams (B) and time course of direct simulation (D) for 3→1 transition.

The transitions between Shifts 2 and 3 share some similarities with those between Shifts 1 and 3. In particular, the switch from Shift 2 to Shift 3 is qualitatively similar to that from Shift 1 to Shift 3, independent of the lux level. This is because the initial value used to compute the reentrainment does not lie near the unstable fixed point in any of the three cases. The ensuing cobweb diagrams and time courses (not shown) are qualitatively similar. However at low light intensity (*I* = 100), the transition from Shift 2 to Shift 3 takes longer than the transition from 3 to 2. This stands in contrast to what was observed between Shifts 1 and 3, where the 3 to 1 transition took longer, due to the initial value lying very close to the unstable fixed point of the map. Figure 8A explains this difference: for the 3 to 2 transition, which is an 8-hour shift of the sleep-wake schedule (rather than the 9-hour shift experienced during the 3 to 1 transition), the initial value *x*_0_ = 1 is not particularly close to the unstable fixed point of the map; see Fig. 8B for the associated time course. The 2 to 3 transition also takes longer than 3 to 2 for the *I* = 1000 and multi-lux cases, similar to what was observed between Shifts 1 and 3. Here, however, the transition from Shift 3 to 2 in the multi-lux case now displays an even shorter entrainment time than the *I* = 1000 lux case. The reason for this is readily discerned from the entrainment maps which show that the unstable fixed point of the multi-lux map (Fig. 8E) lies closer to *x*_0_ = 1 than the unstable fixed point of the *I* = 1000 lux map (Fig. 8C). As before, the proximity of the initial condition to the unstable fixed point for the high- or multi-lux case provides access to the shortcut of the phaseless set, leading to shorter entrainment times. Here the amplitude suppression is greater for the multi-lux case than the 1000 lux case (compare Figs. 8F and D) leading to larger phase changes per iterate and faster entrainment.

**Figure 8:**
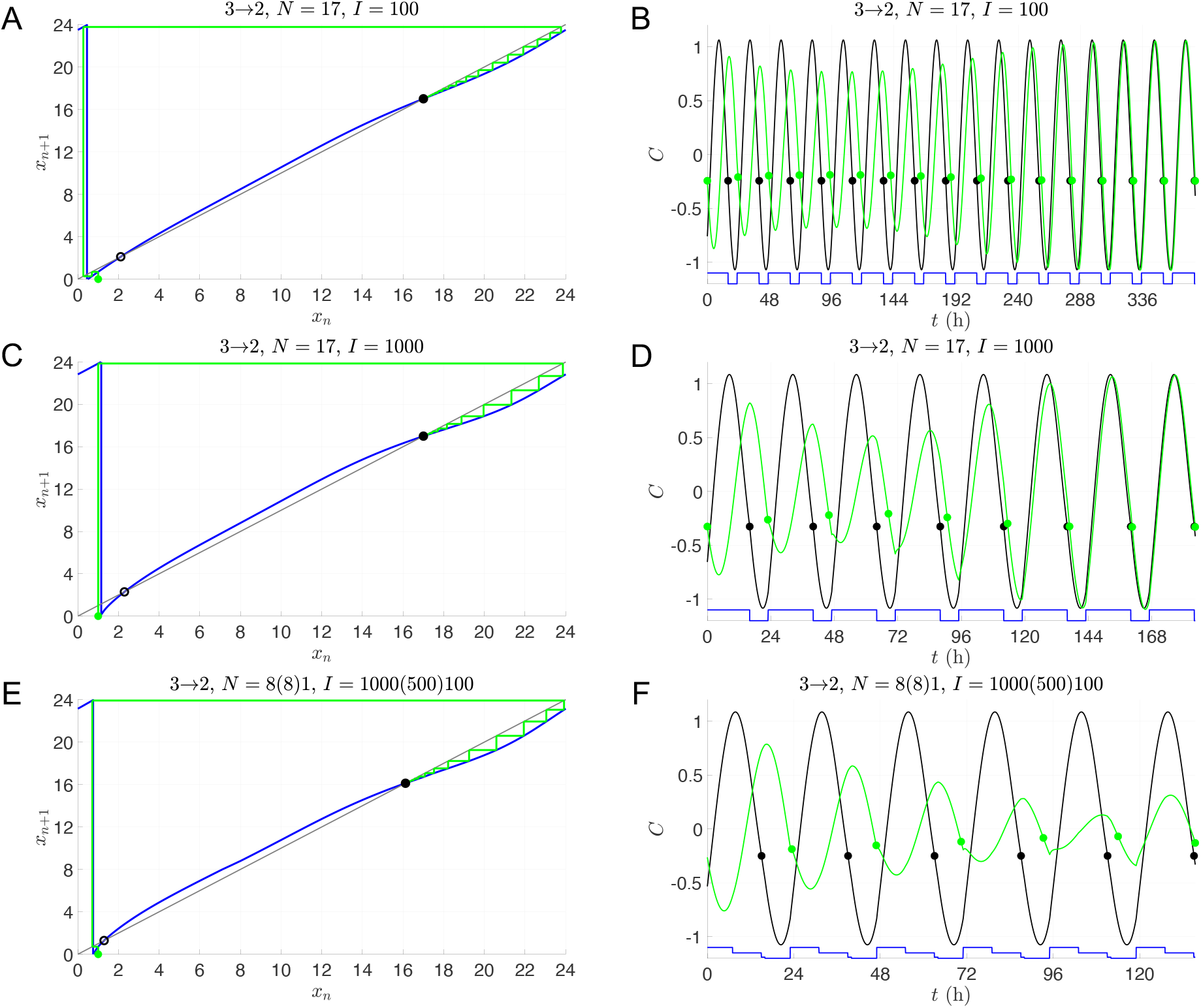
Dynamics of transitions from 3rd to 2nd shift with single- and multi-lux LD protocols. Entrainment maps and cobweb diagrams (A,C,E) and time courses of direct simulations (B,D,F) for *I* = 100 (A-B), *I* = 1000 (C-D), and multi-lux (E-F) LD protocols. Note that a different time scale is shown for each time course.

### 3.3 Social jet lag

Social jet lag can take on many different forms, for example when people stay up late on the weekends, disrupting their normal light exposure patterns. Teenagers also experience a form of social jet lag when they don’t sleep enough during weekdays and compensate by sleeping significantly more on weekends. In both of these cases, individuals do not entrain to the 24-hour LD cycle. Instead, their circadian rhythm displays a complicated periodic pattern that lasts the entire seven-day week. Here we shall focus on social jet lag in teenagers, who typically have a longer intrinsic period than adults [25], and illustrate how the entrainment map can be used to understand their circadian dynamics.

Consider a teenager with a circadian clock of *τ*_*c*_ = 24.6 living under *N* = 17 : 1(12)4, *I* = 100(1000)100. Following this light protocol for an entire week results in entrainment to the LD cycle, the phase of which we can determine by using a single entrainment map. This LD-entrained solution has CBT_min_ occurring at roughly 4AM. Choose a Poincaré section at the plane passing through the 10PM location along this LD-entrained solution. Now imagine that the teenager stays up three hours later on Friday and Saturday evening, but then sleeps three hours later the subsequent mornings. They would no longer entrain to the 24-hour LD cycle but instead their circadian rhythm converges to a complicated 7-day cycle; see Fig. 9A where the red dots indicate that the time of CBT_min_ varies over the week. Observe that the teenager follows the normal schedule of 11PM sleep and 6AM wake only Sunday night through Friday morning, followed by 2AM sleep and 9AM wake up Friday (late) night through Sunday morning. As a result, there are now two different *N* = 17 maps to consider. In addition to the Monday through Friday *N* = 17 : 1(12)4, *I* = 100(1000)100 schedule, on Saturday and Sunday the individual is exposed to *N* = 17 : 10(7), *I* = 1000(100). Figure 9B superimposes these two maps (Monday-Friday map in blue, and Saturday-Sunday in green) and shows the generalized cobweb procedure that produces a stable periodic orbit with this set of maps. The sequence of iterates of the generalized cobweb procedure begins with an initial condition *x*_0_ taken on the section at Thursday night and is shown in Table 1.

**Table 1:** The values *x*_*n*_, *n* = 0, …, 7 refer to the phase of light each time the trajectory crosses the Poincaré section. The notation Π refers to the application of the entrainment map to a particular iterate *x*_*n*_ (or a translation of that iterate as in the 2nd and 4th rows).

| Day | Iterate | $N$ | $I$ |
| --- | --- | --- | --- |
| F | $x_1 = \Pi(x_0)$ | 17 : 1(12)4 | 100(1000)100 |
| Sa | $x_2 = \Pi(x_1 - 3)$ | 17 : 10(7) | 1000(100) |
| Su | $x_3 = \Pi(x_2)$ | 17 : 10(7) | 1000(100) |
| M | $x_4 = \Pi(x_3 + 3)$ | 17 : 1(12)4 | 100(1000)100 |
| Tu | $x_5 = \Pi(x_4)$ | 17 : 1(12)4 | 100(1000)100 |
| W | $x_6 = \Pi(x_5)$ | 17 : 1(12)4 | 100(1000)100 |
| Th | $x_7 = \Pi(x_6)$ | 17 : 1(12)4 | 100(1000)100 |

**Figure 9:**
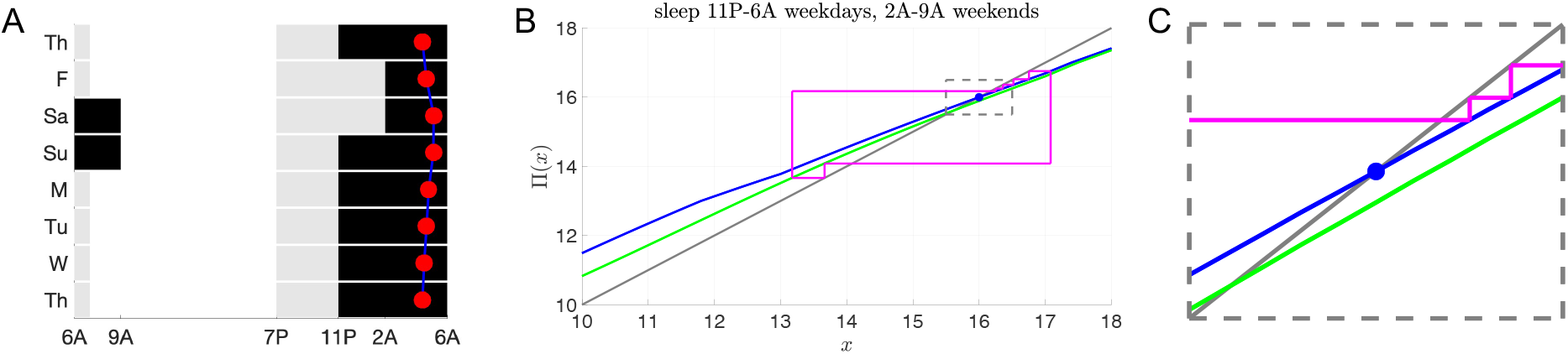
Social jet lag due to a shift in sleep and wake times on weekends. *A:* Schematic of the LD protocol for a teenager (*τ*_*c*_ = 24.6) that sleeps (black shading, 0 lux) from 11PM to 7AM on weekdays and from 2AM to 9AM on weekends. Gray shading indicates indoor light (100 lux) from 7PM to 11PM and white shading indicates outdoor light (*I* = 1000) from 7AM to 7PM. Red dots indicate the time of CBT_*min*_ in direct simulations. *B:* Entrainment maps and generalized cobweb diagram for the sleep/wake schedule shown in (A). Blue curve is the weekday map *N* = 1(12)4, *I* = 100(1000)100 with a stable fixed point at *x*_*s*_ = 16 (solid blue dot). Green curve is the weekend map *N* = 10(7), *I* = 1000(100). Cobweb iterates (magenta) converge to a 7-day periodic orbit. Weekday iterates outside of the dashed gray box are more than 0.5 hours away from *x*_*s*_ and are considered to be misaligned. *C:* Magnification of the maps and cobweb iterates inside the dashed gray box shown in panel B.

Note that because of the change of light exposure on certain days, there are horizontal shifts of three hours of the trajectory between Friday and Saturday and between Sunday and Monday. The shift three hours to the left on Saturday reflects the delay of three hours in the wake time on weekends. The shift three hours to the right on Monday is the advance back to the earlier weekday wake time. One measure of the extent of circadian misalignment is to track the values of the iterates *x*_*i*_ corresponding to the weekdays to determine how many of them lie within 30 minutes of the stable fixed point *x*_*s*_ of a fully entrained individual. In the case shown in Fig. 9B, the Monday iterate does not fall within this window.

Having described how we use multiple maps over the 7-day week, let us now address a specific concern for high school students. Many do not sleep the recommended 8 to 10 hours per night during the week; instead, they sleep less due to staying up late trying to finish homework. Then, they try to compensate by sleeping significantly longer on the weekends [26, 27]. Thus these teenagers suffer from the combination of social jet lag and sleep deprivation. Students in different countries handle this problem in different ways. For example, Carskadon [26] notes that Korean high school students compensate by going to sleep earlier on weekends than students from North America. We now demonstrate, using entrainment maps, that this strategy of going to sleep earlier on the weekends is indeed advantageous in minimizing circadian misalignment as long as the sleep time is not too early.

Consider an 11th or 12th grader who follows a 12AM to 6AM sleep schedule Sunday night through Friday morning, which mirrors the average weekday sleep and wake times reported by [26]. We investigated the extent of their circadian misalignment under different weekend sleep protocols of 10 hours (Fig. 10). The rows correspond to progressively earlier weekend sleep onset times from 2AM to 8PM. The first column shows the LD protocol (gray 100 lux, white 1000 lux, black 0 lux). The red dot signifies the time of CBT_min_. Note there is minimal deviation in CBT_min_ for 10PM sleep (panel G). Column 2 shows corresponding cobweb diagrams. The weekday (blue) and weekend (green) maps have different fixed points. During the weekdays, the iterates approach the stable fixed point of the blue map, while on the weekend they approach those of the green map. The magnitude of the horizontal shifts progressively get smaller as the wake onset time shifts earlier, with the earliest sleep time of 8PM in panel K having no horizontal shifting of iterates since the weekend wake time of 6AM is the same as the weekday wake time. The dynamics within the dotted rectangle of the weekday iterates that fall within 0.5 hours of the stable fixed point value of *x*_*s*_ = 16 are shown in column 3 (1, 3, 5, and 4 iterates, respectively). From these maps, it is clear that sleeping at 10PM on weekends leads to minimal changes of iterate values between Monday and Friday and minimal changes from Saturday to Sunday (Panel H). To understand why, observe that the stable fixed point of the *N* = 18 weekday map at *x* = 16 is roughly two hours away from the fixed points of the *N* = 14 weekend maps. This difference between the value of the fixed points counteracts the two-hour horizontal shifts caused by waking up at 8AM.

**Figure 10:**
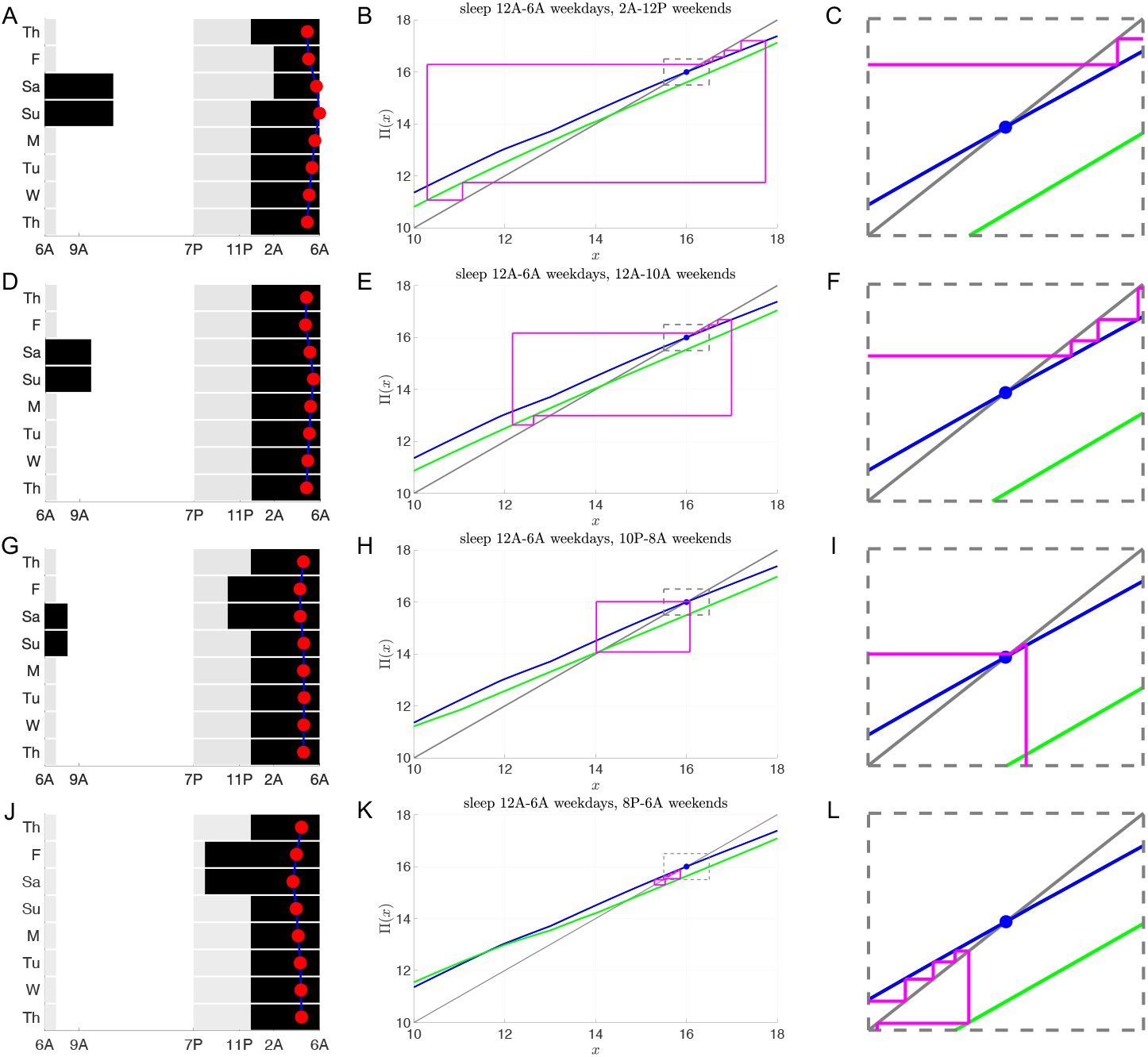
Social jet lag with sleep deprivation during the week and recovery sleep on the weekends. Schematics of LD protocols (left column) and entrainment maps/cobweb diagrams (middle and right columns) following the same format as Fig. 9 for a teenager (*τ*_*c*_ = 24.6) sleeping 6 hours (from 12AM to 6AM) on weekdays and 10 hours on weekends. Weekday map is for *N* = 1(12)5, *I* = 100(1000)100. *A-C:* Weekend sleep begins at 2AM and weekend map is *N* = 7(7), *I* = 1000(100). *D-F:* Weekend sleep begins at 12AM and weekend map is *N* = 9(5), *I* = 1000(100). *G-I:* Weekend sleep begins at 10PM and weekend map is *N* = 11(3), *I* = 1000(100). *J-L:* Weekend sleep begins at 8PM and weekend map is *N* = 1(12)1, *I* = 100(1000)100.

The above observation about the distance between fixed points suggests a strategy to minimize circadian mis-alignment by choosing sleep and wake onset times appropriately. Let *N*_*wd*_ and *N*_*we*_ signify the photoperiods on the weekdays and weekends, respectively. Let Δ*x*_*s*_ denote the distance in the fixed points of the respective maps. We have observed over a wide set of photoperiods that Δ*x*_*s*_ *≈* (*N*_*wd*_ − *N*_*we*_)*/*2. Thus we propose the following “rule of thumb” to minimize misalignment: shift both the sleep onset time forward and the wake onset time backward by (*N*_*wd*_ − *N*_*we*_)*/*2. To quantify the effect of such a strategy relative to other sleep and wake onset times, we plot the *L*^1^ norm of the weekday iterate distances from *x*_*s*_ = 16 for *N*_*wd*_ = 18 and *N*_*we*_ = 8 (red), 10 (blue), 12 (black) for different sleep onset times (Fig. 11). Each graph is V-shaped with a local minimum occurring when sleep onset occurs (*N*_*wd*_ − *Nwe*)*/*2 before 12AM, which is the weekday sleep onset time. Thus, either going to sleep overly early or staying up too late contributes to misalignment. The results suggest that given a weekday sleep pattern, a specific weekend sleep protocol can be chosen to minimize misalignment while still catching up on missed sleep. In particular, the delay in wake up time of the weekend map should equal the distance between the fixed points of the weekday and weekend maps. In turn this delay in wake up time accounts for half of the difference in the weekday and weekend photoperiods, thus suggesting the optimal time to go to sleep.

**Figure 11:**
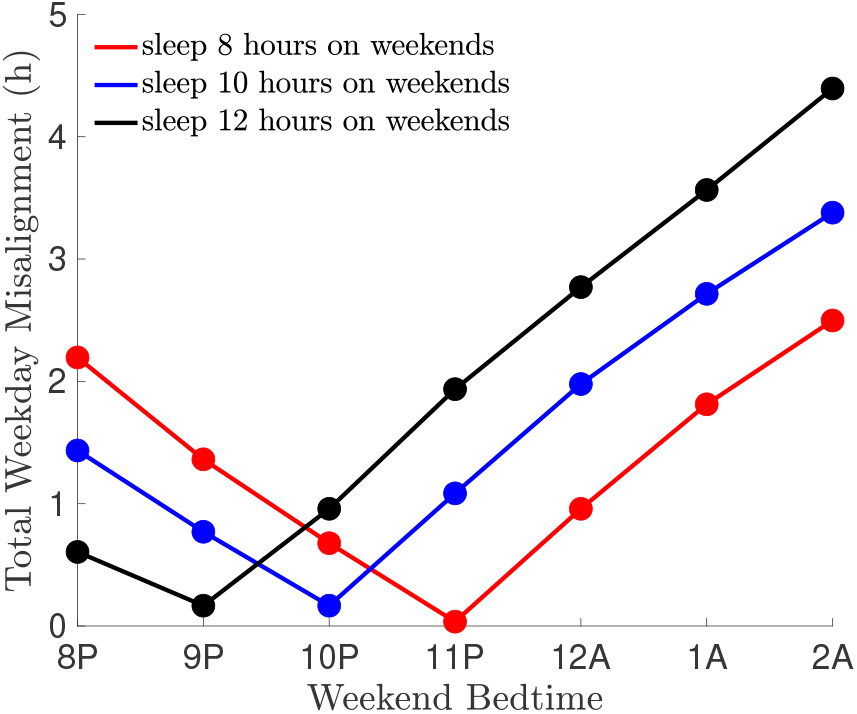
Degree of misalignment due to social jet lag as a function of weekend bedtime for various amounts of weekend recovery sleep. All curves correspond to sleeping 6 hours on weekdays (*N*_*wd*_ = 18) and 8 (red; *N*_*we*_ = 16), 10 (blue; *N*_*we*_ = 14), or 12 (black; *N*_*we*_ = 12) hours on weekends. The local minimum in all cases occurs at a weekend bedtime of 12 − (*N*_*wd*_ − *N*_*we*_)*/*2.

## 4 Discussion

In this paper, we have investigated non-entrainment in specific circumstances that arise due to either biological reasons (non-24-hour sleep wake disorder) or societal pressures (shift work and social jet lag). Entrainment of circadian rhythms to a 24-hour light-dark cycle has been empirically demonstrated to be of importance for the health and well-being of humans. The vast majority of us are able to achieve this entrainment throughout our lives, even as our endogenous circadian oscillations slowly change over our lifespan. We do so in the face of an ever changing social landscape, here taken to mean changing LD exposure patterns on a day to day basis or throughout the week. In short, our circadian system is quite robust to perturbations. There are, however, several circumstances where we entrain at a phase that is not ideal or lose the ability to entrain to a 24-hour LD cycle entirely. Such circadian misalignment can have negative consequences on a person’s physical or mental health, behavior, and performance.

### 4.1 Main findings and strategies for counteracting circadian misalignment

#### Non-24-hour sleep-wake disorder

This disorder can be characterized as an inability to entrain to the 24-hour LD cycle. Using entrainment maps [17, 18], we showed that the non-entrainment corresponds to the lack of existence of a stable fixed point of the map. The dynamics obtained by cobwebbing such maps are consistent with the relapsing and remitting nature of the sleep issues observed in non-24 patients. Specifically, we found that several consecutive map iterates occur near to where the fixed point would be located at the point of saddle-node bifurcation and exhibit small phase advances or delays, a phenomenon referred to as slow passage through the ghost of a fixed point [28]. This dynamical phenomenon corresponds to several days of nearly entrained behavior and a lack of sleep issues in non-24 patients. These small phase shifts are followed by a series of iterates with large phase advances or delays that progress further and further away from the ghost of the fixed point until the phase of CBT_min_ progresses through the entire 24-hour cycle. These iterates correspond to many days of completely unentrained behavior and sleep issues in non-24 patients. Understanding what led to non-entrainment subsequently allowed us to devise strategies to induce the existence of a stable fixed point. For those with a slow endogenous clock, we showed that bright light therapy early in the day induces a stable fixed point corresponding to CBT_min_ occurring during sleep; bright light therapy administered later in the day does not induce entrainment. This finding is consistent with the suggestions made to patients to use bright light therapy upon awakening [8, 19]. Surprisingly perhaps, we showed that early morning bright light therapy for patients with fast body clocks is not therapeutic. While this therapy produces entrainment in such individuals, it does so at a phase that leads to CBT_min_ occurring during midday. Our analysis suggests that for individuals with a fast body clock, bright light therapy should be administered in the early evening. This results in entrainment at a phase where CBT_min_ occurs during normal sleep hours. The results found here are consistent with the overall theme that individualized treatments for circadian disorders may lead to more efficacious results and better alignment of the circadian cycle [29].

#### Shift work

We addressed whether workers would experience excessive circadian misalignment during the transition between working on different shifts (day, evening, or night). A primary concern here is that CBT_min_ might occur either during work hours or during the drive to or from work, which might signal excessive drowsiness during these times. First, we showed that the reentraiment times when adjusting to a new work shift are asymmetric and highly dependent on light intensity levels. At low light intensity, the switch from the night shift to the day shift has the longest reentrainment time. Conversely, in the presence of more realistic and higher intensity light protocols, reentrainmnent for the night to day shift transition is substantially faster than the transition from day to night shift. This fast reentraiment can be explained in mathematical terms due to the existence of shortcuts in phase space that allow the circadian oscillator to access a region known as the phaseless set [20]. We [18] and others [30, 31] have shown that passage of trajectories through the phaseless set are characterized by amplitude suppression as the circadian oscillator reentrains. The association between the amplitude and a period of an oscillator, referred to as “twist”, facilitates short reentrainment times in a minimal model of the molecular clock [32]. Creaser et al [31] have shown that reentrainment times following transitions between day and night shift work are organized by invariant manifolds of entrained periodic orbits. Specifically, the shortcut in phase space that enables fast reeentrainment when transitioning from night shift to day shift at high light intensity is associated with the strong stable manifold of the stable entrained limit cycle. In contrast, at low light intensity, there also exists an unstable saddle entrained limit cycle, and trajectories that begin near the stable manifold of this unstable limit cycle take a long time to reentrain following transitions from night to day shift. Overall the asymmetry in reentrainment times in shift work is similar to the east-west asymmetry experienced in recovery from jet lag. In this situation, individuals with a normal body clock (≈24.2 hours) take longer to recover from travel to the east, rather than to the west. In the shift work scenario, rotating from 1st to 3rd shift is analogous to traveling west, and indeed we show it takes less time to reentrain than when rotating from 3rd to 1st shift (analogous to traveling east) at low light levels (100 lux). However, at higher light levels (1000 lux and multi-lux), the asymmetry is reversed due to the phaseless set phenomenon described above.

#### Social jet lag

In the context of teenagers who engage in catch up sleep on the weekends, we found that the timing of sleep and wake onset were critical factors in minimizing circadian misalignment. Our results suggest that simply sleeping more on the weekends is not an optimal strategy. A recent study has shown that for people with short sleep duration during weekdays, weekend recovery sleep reduces mortality rate compared to getting no catch-up sleep [33]. However, their study did not assess how sleep timing affects this association [11]. We proposed a rule of thumb for reducing weekday circadian misalignment: move the sleep onset time earlier by half the difference in the photoperiods of weekday versus weekend light exposure, while moving the wake onset time back by this same amount. Following this strategy allows individuals to counteract the changes in photoperiod experienced between weekday and weekend light exposure. In previous work [18], we had suggested that changes in photoperiod resulting from north-south travel (for example between North and South America) could induce jet lag even when there is no change in time zone. Furthermore, we found that the effect of the change in photoperiod due to north-south travel could either counteract or amplify jet lag caused by east-west travel. In that case however, the change in photoperiod is “permanent” in that the individual eventually reentrains to the new photoperiod. In the case of social jet lag considered here, teenagers have just a few days (the weekend) where light is delivered on the shorter photoperiod, followed by more days at the longer photoperiod (weekdays), and then the situation repeats. In this sense, the change in photoperiod is “transient”. Empirical studies have shown that teenagers from different countries choose different strategies for weekend sleep [26]. The results presented here suggest that with an appropriate choice of earlier weekend sleep and later wake onset times, teenagers can dramatically reduce their circadian misalignment and thereby improve their performance in school.

### 4.2 Relation to prior work

As there is a vast empirical and modeling literature related to entrainment of circadian rhythms [34–38], we shall focus on just a few of the works that are related to our current study. Papatsimpa et al [39] use the Kronauer model to study how modern lifestyles and the effects of different light levels lead to a shift towards more evening chronotypes. In attempting to minimize social jet lag due to dim indoor work lights, Papatsimpa and Linnartz [40] design personalized lighting schedules based on minimizing spontaneous versus alarm wake up times. Julius et al [41] study entrainment of the Kronauer model to obtain the fastest possible reentrainment in the context of jet lag. These latter two papers use optimal control theory. Based on collected data, they train algorithms that suggest, sometimes on an hourly basis, what light intensity level individuals should be exposed to in order to minimize misalignment. Our work here, while seemingly less individualized, suggests strategies for circadian management using a mechanistic approach. In particular, we showed that for groups of individuals of different chronotypes, there are specific interventions that can lead to entrainment (e.g. early morning or late evening bright light therapy to combat non-24-hour sleep-wake disorder) or appropriate adjustment of sleep/wake onset (e.g. for teenagers with slower body clocks who engage in compensatory sleep on the weekends). Determining the endogenous circadian clock speed of individuals remains a challenging problem to overcome at scale. The cost and time of invasive measures of data collections such as regular sampling of blood is prohibitive. Recent work of Cheng et al [42] suggests that wrist-worn actigraphy combined with photometry can estimate circadian phase. This is a promising approach as it will allow for greater individualization of treatment protocols to reduce circadian misalignment.

Our work did not contain a model with an explicit sleep component. As such we did not address the question of whether sleep timings, controlled in part by the sleep homeostat and by circadian forcing, are matched to circadian rhythmicity. There are several models that do study questions related to how these two coupled oscillatory systems interact. In the context of shift work, Postnova et al [43] suggest that the intensity of shift lights can be reduced and still produce a transition from day to night shift work without excessive sleepiness. Kim et al [44] extended the Potsnova model to include the effect of circadian drive onto orexin neurons, which in turn affect the sleep/wake system. Their findings show that both a lack of light and imbalance of the orexin system can lead to seasonal affective disorder and lack of entrainment. Consistent with our findings here, they show that bright light therapy administered early in the day can mitigate this misalignment over a wider range of parameters than can such therapy later in the day. Skeldon et al [45] model how self-selected light schedules affect circadian rhythms. Their work suggests that minimizing the amount of artificial light in the evening is an effective way to align sleep and circadian rhythms. A potential area of future research for us would be to couple the entrainment map with a map that predicts sleep-wake onset times as in [46].

### 4.3 Conclusion

Circadian misalignment can lead to detrimental outcomes for individuals and the societies in which they live. Mathematical models can provide insight into the underlying mechanisms that cause misalignment as well as suggest strategies to combat it. In this work, we have used a mathematical and computational analysis of entrainment maps in three specific cases that go beyond the normal limits of circadian entrainment. Depending on the context, we showed how to either induce entrainment, minimize reentrainment times, or minimize the extent of circadian misalignment. Our hope is that our methodology and findings can be combined with complementary approaches to develop more individualized approaches to circadian management.

## Data Availability

Code will be provided upon request.

## Data Availability

Code will be provided upon request.

## Data Availability

Code will be provided upon request.

## 5 Acknowledgments

This material is based upon work supported by the National Science Foundation under grant no. DMS 1555237 and the U.S. Army Research Office under grant no. W911NF-16-1-0584. We are grateful for fruitful conversations with Dr Kyle Wedgwood and Dr Jen Creaser.

